# Prevalence Threshold and Temporal Interpretation of Screening Tests: The Example of the SARS-CoV-2 (COVID-19) Pandemic

**DOI:** 10.1101/2020.05.17.20104927

**Authors:** Jacques Balayla, Ariane Lasry, Yaron Gil, Alexander Volodarsky-Perel

**Affiliations:** Department of Obstetrics and Gynecology, McGill University, Montreal, Quebec, Canada

**Keywords:** screening curve, positive predictive value, prevalence threshold, COVID-19, Bayes’ Theorem

## Abstract

The curvilinear relationship between a screening test’s positive predictive value (PPV) and its target disease prevalence is proportional. In consequence, there is an inflection point of maximum curvature in the screening curve defined as a function of the sensitivity (*a*) and specificity (*b*) beyond which the rate of change of a test’s PPV declines sharply relative to disease prevalence (*ϕ*). Herein, we demonstrate a mathematical model exploring this phenomenon and define the prevalence threshold point (*ϕ_e_*) where this change occurs as:

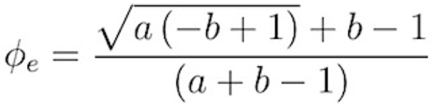

Understanding where this prevalence point lies in the curve has important implications for the interpretation of test results, the administration of healthcare systems, the implementation of public health measures, and in cases of pandemics like SARS-CoV-2, the functioning of society at large. To illustrate the methods herein described, we provide the example of the screening strategies used in the SARS-CoV-2 (COVID-19) pandemic, and calculate the prevalence threshold statistic of different tests available today. This concept can help contextualize the validity of a screening test in real time, thereby enhancing our understanding of the dynamics of the current pandemic.

## Introduction

The novel SARS-CoV-2 (COVID-19) has reached over 4 million confirmed cases worldwide; in the United States alone, the death toll totals close to 90,000 to date [1]. Despite efforts to contain its spread, the number of confirmed cases has continued to rise. These estimations have been largely based on polymerase chain reaction (PCR) detection of actively replicating viral material, found only among individuals who are actively infected at the time of testing. Those who recover from the virus without having been tested during the window of viral shedding are not included in prevalence estimates; neither are asymptomatic individuals, roughly approximated as 18 to 30% of those infected [2, 3]. Current estimates thus understate the true prevalence of COVID-19 by failing to include the aforementioned groups. Whereas accurate prevalence estimates are needed to inform public health measures, screening methods that depend on actively replicating viral organisms will become less reliable as recovery proceeds and the resulting prevalence of active infection among the population tapers. Bayes theorem thus demonstrates the point beyond which the positive predictive value (PPV) of currently employed PCR screening will decline as the curve ‘flattens’ over time. In contrast, serologic testing for COVID-19 antibodies persist in plasma well beyond the period of active infection; as recovery from active infection increases, so too will the cumulative prevalence of seropositivity among the population over time. Herein, we use differential equations to assess the geometry of screening curves and aim to describe the prevalence threshold point beyond which the PPV of various COVID-19 screening tests declines most acutely. Though this example is specific to the COVID-19 pandemic, the methods herein described apply to all screening tests whose sensitivity and specificity parameters are known.

### Validity of Screening Tests

The validity of a screening test is defined as the ability to correctly delineate individuals who have a given disease or condition from those who do not. The following four parameters are used to assess the validity of screening tests: sensitivity *a*, specificity *b*, positive predictive value *ϕ*, and negative predictive value σ. Sensitivity refers to the proportion of people with a given disease who test positive for said disease, also termed the true positive rate. Specificity, also termed true negative rate, refers to the proportion of people without said disease who indeed test negative. Sensitivity and specificity are properties inherent to the screening test itself and are unaffected by the prevalence of disease in a given population. On the other hand, the positive predictive value is defined as the percentage of patients with a positive test that do in fact have the disease, and conversely, the negative predictive value refers to the percentage of patients with a negative test that do not have the disease. These two parameters are dependent upon the prevalence of disease being studied. Using Bayes’ theorem, we can derive the following equation expressing positive predictive value *p*(*ϕ*) as a function of disease prevalence *ϕ*.

**Equation 1. *The Screening Curve Equation***

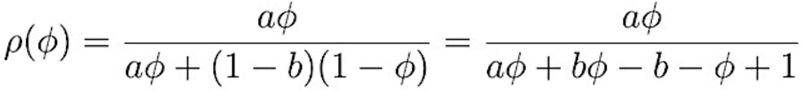

where:

*ϕ* = prevalence, *p*(*ϕ*) = positive predictive value, *a* = sensitivity and *b* = specificity

### Prevalence Threshold of a Screening Test

Bayes’ theorem describes the probability of an event, based on prior knowledge of conditions that might be related to the event. The relationship between a screening test’s positive predictive value, *p*(*ϕ*), and its target disease prevalence *ϕ* proportional - though not linear in all but one case where the sum of sensitivity and specificity equals one. From this curvilinear relationship, as stipulated in Figure 1, we can derive the prevalence threshold *ϕ_e_* at which the sharp inflection point in the screening curve occurs, as depicted by the following equation (derivation available as a supplement).

**Equation 2. *The Prevalence Threshold Equation***

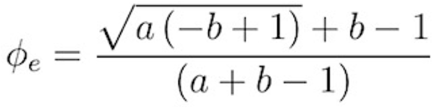

where:

*ϕ_e_* = prevalence threshold, *a* = sensitivity and *b* = specificity

### Nasopharyngeal Swab RT-PCR Screening Test for COVID-19

The Food and Drug Administration (FDA) published the analytical sensitivity and specificity of the currently employed COVID-19 reverse transcription polymerase chain reaction (RT-PCR) test [4], The limit of detection (LoD) for the minimum detectable concentration of SARS-CoV-2 genome (ie. sensitivity) in-vitro was set at 95%. Additionally, the COVID-19 RT-PCR was tested against 30 respiratory microorganisms, which yielded a specificity nearing 100%. While the above are *analytical* properties of the COVID-19 RT-PCR test, estimates of its *clinical* properties vary significantly between studies – particularly since some margin of error may result from improper sampling by health care workers (wrong angle, too fast to withdraw and swab, etc.), varying viral load and the subclinical stage when screening is carried out (colonization, incubation, prodrome or acute infection). We will therefore use the FDA’s published estimates for simplicity, where *ϕ_e_* = prevalence threshold for detection of COVID-19, *a =* sensitivity = 0.95, *b* = specificity = 0.99 and *a + b =* 1.94. Thus, as per equation 2, the prevalence threshold for RT-PCR detection of COVID-19 is calculated as follows:

**Equation 3. *Prevalence threshold for the COVID-19 RT-PCR screening test***

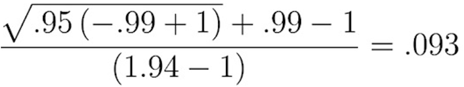

In graphic form, the screening curve depicts the PPV as a function of prevalence as such:

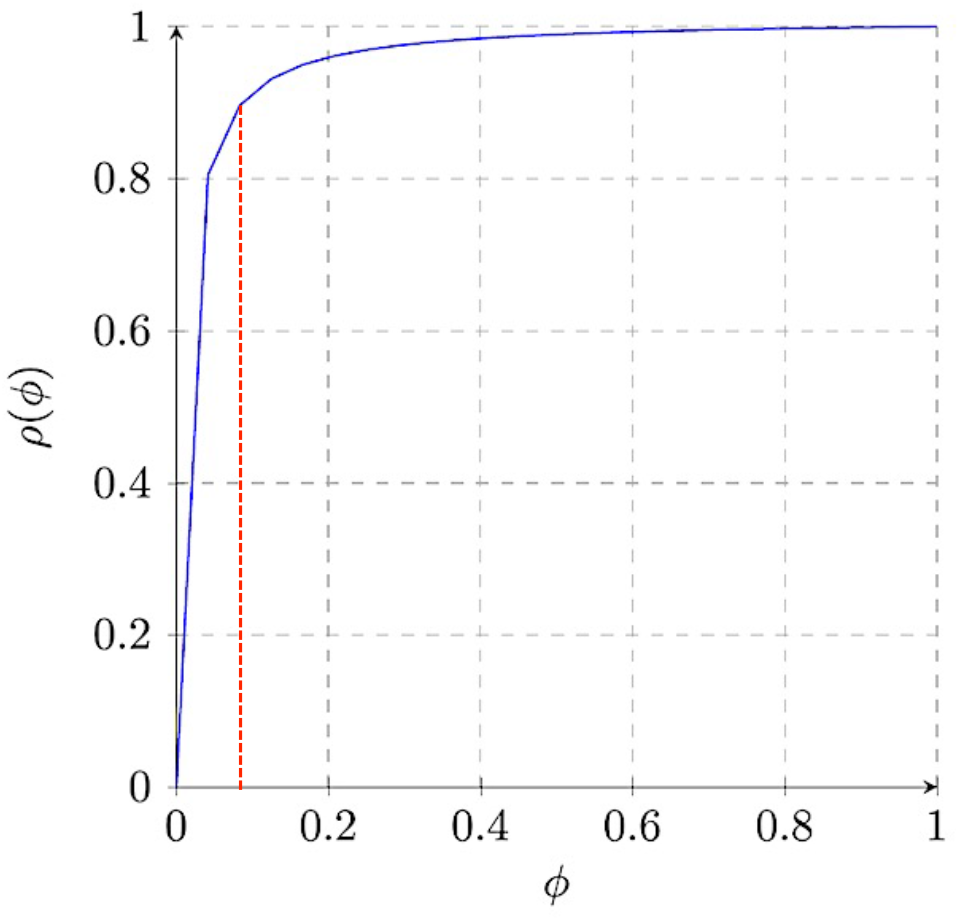

The vertical line in red represents the prevalence threshold *ϕ_e_* at 0.093 (9.3%).

In clinical practice, the sensitivity and specificity of the COVID-19 RT-PCR test are likely lower than those estimated analytically by the FDA; the prevalence threshold for detection that we have estimated above is thus likely conservative. Nonetheless, the implication is such that below a prevalence of 9.3% actively replicating cases of COVID-19 in the population, the PPV of RT-PCR testing declines almost exponentially. In other words, we would expect a sharp increase in the number of false positive screening tests, in turn falsely increasing the estimated prevalence of disease. Even with a conservative estimation of the prevalence threshold, there are potentially significant health, social, and economic implications of false positive screening tests.

### Serology Testing as a Public Health Tool

Serology testing for SARS-CoV-2 involves blood-based testing for antibodies to COVID-19. This screening method identifies all groups of individuals sub-acutely infected or recovered from COVID-19, including those who may be asymptomatic at the time of testing. Use of this tool can thus provide public health officials with a more reliable estimation of the spread of COVID-19 and its cumulative prevalence among different populations over time. Furthermore, this information can bring forth attempts at a better understanding of disease transmission and immunity, which regarding COVID-19, is largely uncertain up until this point.

The Johns Hopkins Center for Health Security recently released a report outlining the sensitivity and specificity of various serology tests approved for diagnostic use in the United States [5]. These are summarized in Table 1, with prevalence thresholds estimated for each test. In contrast to RT-PCR screening, COVID-19 testing with serology can delineate immune individuals at a prevalence threshold as low as 4.3%. As the seropositive rates continue to increase, we deduce that above a prevalence of 4.3%, a positive COVID-19 IgG screen can be reliably accepted as a true positive. In contrast, the nasal RT-PCR test is most reliable when over 9.3% of the population has actively replicating virus at any given time – a value thankfully thus far not reached, even after accounting for an excess positive cases that are not tested.

**Table 1.**
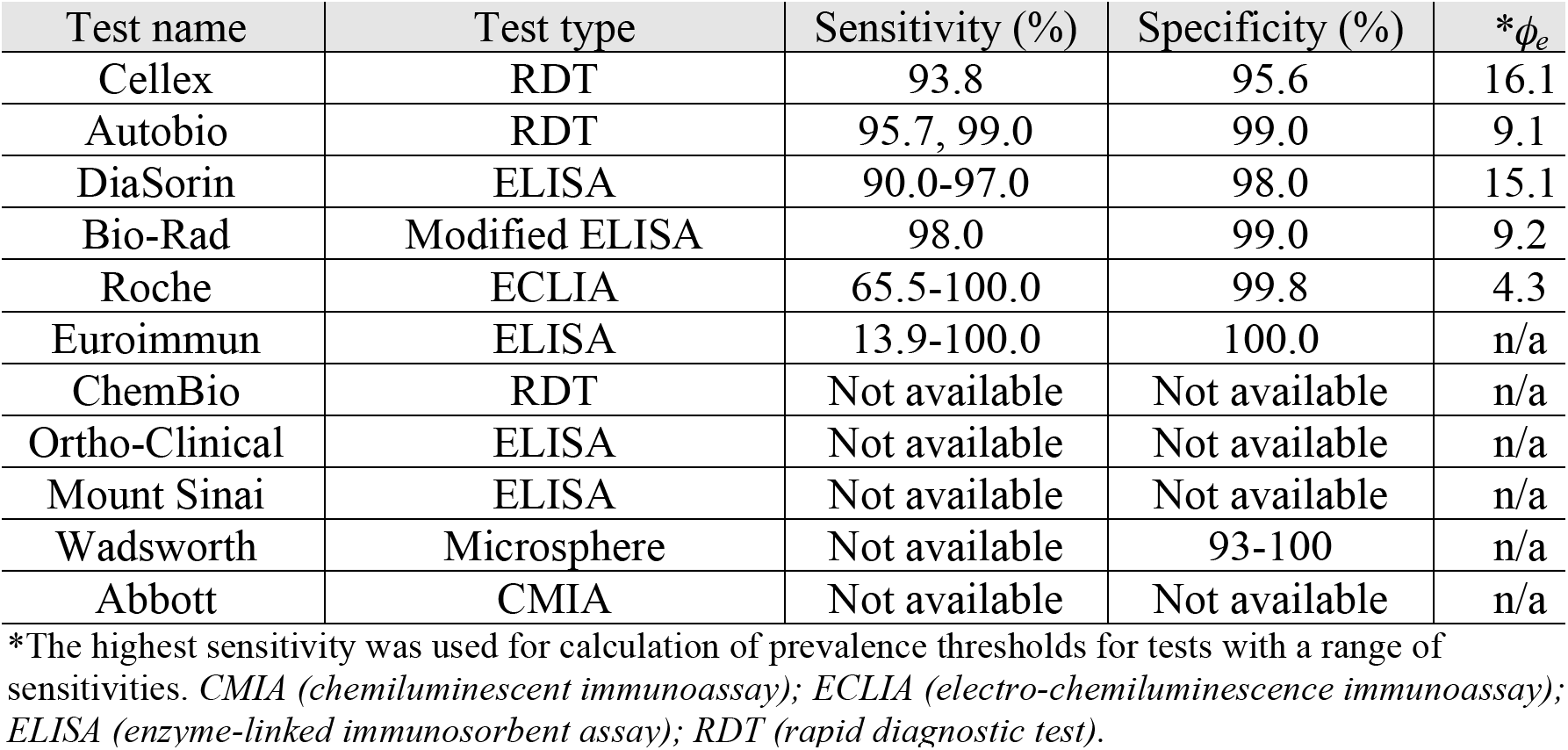
Prevalence thresholds (ϕ_e_) for different serologic tests available.

Several countries have already started employing serology-based testing, either for research purposes or to grant ‘immunity certificates’ to those in whom antibodies to COVID-19 are found [6]. However, we must bear in mind that just as with RT-PCR testing, serologic testing has inherent limitations related to the lag between acute infection and the development of IgM and IgG antibodies as well. In the United States, the Centers for Disease Control and Prevention (CDC) recently released a COVID-19 Serology Surveillance Strategy [7]. This strategy aims to employ serology testing, termed ‘seroprevalence,’ at the large-scale (ie. highly impacted areas such as New York and Washington), the community-scale (with systematic selection of participants) and the small-scale (specific subgroups, eg. healthcare workers). From the reasoning above, we conclude that the best screening method to assess for acute infectivity, the need for isolation, and the ensuing social and economic repercussions, is nasal swab RT-PCR for actively replicating virus. In contrast, the most reliable screening method to assess for disease prevalence and burden of disease is antibody testing via serology – assuming enough time has lapsed since the acute infectious episode.

### Note on Rapid Diagnostic Testing

In an effort to expedite testing speed and availability, rapid diagnostic tests (RDT) have been developed for point-of-care use [8-17]. These tests rely on serological markers against SARS-CoV-2, either IgM or IgG, for diagnosis of acute COVID-19 infection. While the FDA has approved various RDTs for use under Emergency Use Authorization, the WHO currently does not recommend their use outside of research purposes due to their inconsistent sensitivities [18]. Indeed, concerns over reports of false negative results prompted an FDA news release cautioning the public about the potential inaccuracy of Abbott’s ID NOW point-of-care RDT [13-17, 19]. Further studies with larger sample sizes and clear manufacturer instructions for correct sampling are needed in order to determine whether RDTs are reliable for clinical use.

## Conclusion

The aforementioned values for sensitivity and specificity are likely to change as more samples are obtained. However, their values as stipulated here are used not to endorse a particular test, but to illustrate the concept of prevalence thresholds in the context of the current pandemic. To illustrate the methods herein described, we provide the example of the screening strategies used in the SARS-CoV-2 (COVID-19) pandemic, and calculate the prevalence threshold statistics of different tests available today. This concept can help contextualize the validity of a screening test in real time, thereby enhancing our understanding of the dynamics of the current pandemic. [Identifying where this prevalence point lies in the curve has important implications for the interpretation of test results, the administration of healthcare systems, the implementation of public health measures, and in cases of pandemics like SARS-CoV-2, the functioning of society at large.

## Data Availability

We used publicly available estimates for sensitivities and specificities of different COVID-19 tests, as published by the Center for Health Security at John Hopkins University.

https://www.centerforhealthsecurity.org/resources/COVID-19/serology/Serology-based-tests-for-COVID-19.html

